# Machine Learning with Objective Serum Markers and Algorithmic Deep Learning Computed Tomography Scan Analysis for Classification of Brain Injury

**DOI:** 10.1101/2021.02.13.21250776

**Authors:** Daniel Rafter, Zhuliu Li, Tory Schaaf, Kristen Gault, Maxwell Thorpe, Shivani Venkatesh, Radhika Edpuganti, Tianci Song, Rui Kuang, Uzma Samadani

## Abstract

**Background:** Brain injury is pathophysiologically diverse, with many cases presenting with mixed pathologies. Utilizing objective measures to investigate the pathophysiology of injury would aid in understanding prognosis and targeting therapeutics.

**Objective:** The goal of this study is to develop a traumatic brain injury classification scheme based on open source deep learning computer tomography (CT) analysis and the two serum biomarkers, glial fibrillary acidic protein (GFAP) and ubiquitin carboxy-terminal L1 (UCH-L1).

**Methods:** Machine learning was utilized to develop a novel algorithm capable of classifying the type of brain injury based on a CT scan analysis algorithm and GFAP and UCH-L1 concentrations. Injury was stratified into one of four groups: spontaneous hemorrhage, oxygen deprivation, trauma resulting in vascular injury or high-velocity trauma with negative CT scan.

**Outcomes:** 100 research subjects were enrolled. Using a combination of CT analysis and serum markers, the subjects with CT positive trauma were distinguishable from those with spontaneous hemorrhage, ischemic injury, CT negative trauma and controls with AUCs of 0.96, 0.99., 0.98 and 1.00 respectively. Ischemic injury was distinguishable from CT positive trauma with an AUC of 0.98. All forms of brain injury could be distinguished from controls with AUC = 1.00.

**Discussion:** An open source algorithmic CT scan analysis algorithm and serum biomarkers accurately classified the nature of brain injury across major etiologies. Further implementation of such algorithms and addition of other objective measures will enable better prognostication of injury and improved development of therapeutics.

## Introduction

The objective classification of brain injuries using markers for the heterogeneous pathophysiology indicative of different mechanistic forces of injury remains a challenge. Currently, the primary assessors used to classify brain injuries in an acute care setting are the patient’s medical history, physical examination using measures such as the Glasgow Coma Scale (GCS), and radiographic imaging with computed tomography (CT). However, history and physical examination are impacted by confounding factors such as intoxication, comorbidities, language barriers, cultural norms and examiner biases. Conventionally read CT scans do not provide a quantitative measure of injury.

The diverse pathophysiologic mechanisms that contribute to neurological damage in brain injury can be assessed with serum biomarkers to create specific injury profiles of traumatic and nontraumatic brain injuries.^1^ Glial fibrillary acidic protein (GFAP) and ubiquitin c-terminal hydrolase-L1 (UCH-L1) are serum based biomarkers with neurologic specificity^1,2^ and the potential to improve classification of injury. For example, axonal shearing may result in immediate release of cellular contexts during a traumatic injury, in contrast to a delayed release of cellular contents seen in oxygen deprivation injuries; a result of slower apoptosis.^1,3^ This information is useful in predicting the most effective therapeutic interventions.

The objective of the current study was to create an objective classification scheme for heterogeneous brain injury. UCH-L1 and GFAP concentrations were assessed in the acute period of less than 32 hours after injury in order to differentiate among the following five groups: healthy controls, CT negative TBI with high-velocity trauma (CTN-HVT), oxygen deprivation injuries from cardiac or respiratory arrest (CA/RA), spontaneous hemorrhage, and traumatic injury resulting in a CT scan positive for vascular injury. The 32 hour time point was chosen based on previous work demonstrating GFAP and UCH-L1 concentrations during this time frame can be used to accurately predict CT positive versus negative scans among patients with traumatic brain injuries.^4^ BLAST-CT, an open source, automated deep learning CT analysis algorithm was added to improve accuracy of classification.^5^ We hypothesized that each group would have a different biomarker profile because the mechanisms and magnitude in which these biomarkers are released will vary across groups. Successful identification of the differences in these groups will allow for a rapid, accurate classification schema that will optimize triage and treatment times as well as minimize the need for unnecessary medical procedures and potentially improve long term patient outcome.

## Materials and Methods

Institutional Review Board approval was obtained for prospective enrollment of patients who presented to the emergency department of a level-1 trauma center over a one year period. Subjects were consented if they were able to pass the Galveston Orientation and Amnesia Test (GOAT) and met criteria for providing informed consent. If they were not able to provide informed consent initially, a legally authorized representative was consented and the subject was reconsented once able. Only in situations where the subject expired in the hospital and family or a legally authorized representative could not be contacted was informed consent waived. Informed consent or waiver of consent was obtained prior to data inclusion. Detailed description of study protocol and blood draw analysis can be found in supplemental materials.

Analysis and plotting were performed using MATLAB (R2018b). Only samples with both valid UCH-L1 and GFAP biomarker results were included in the analysis. Biomarker levels below the detectable limit prior to transformation were assigned a value of zero, but included in the analysis. Biomarker concentrations of UCH-L1 and GFAP were taken from the same patient’s blood draw and were log transformed for development of the machine learning algorithm. Biomarker levels above the detectable limit (50,000 pg/ml for GFAP and 20,000 pg/ml for UCH-L1) were included in analysis as equal to the upper limit. Only six GFAP and three UCH-L1 blood draws had concentrations above the detectable limit.

Differences in mean and median raw concentrations between traumatic injury, spontaneous hemorrhage (SpontHem), oxygen deprivation (CA/RA), Computer tomography negative- high-velocity trauma (CTN-HVT) and control groups at all timepoints measured within 32 hours were assessed using the t-test and Wilcoxon rank-sum test. Concentrations were log-transformed to achieve normality prior to the plotting and data analyses.

Classification based on the integrated features of serum biomarkers and CT scan images was performed using a pre-trained deep learning model (BLAST-CT) based on a convolutional neural network.^5^ This algorithm automatically generates the voxel-wise segmentation of 4 lesion-types: intraparenchymal hemorrhage, extra-axial hemorrhage, perilesional edema and intraventricular hemorrhage. BLAST-CT identified the regions of the four lesion-types and quantified their respective volumes by counting the total segmented voxels. The segmentation volumes were normalized by the total head volume in each scan.

## Machine Learning

Support Vector Machine (SVM)^6^ was utilized for the classification of the patient samples in our prediction tasks (see supplementary methods). We performed leave-one-out cross-validation to evaluate the prediction performance of SVM. In the evaluation, we hold out one sample at a time as the test data and treat the rest samples as training data to obtain the SVM model for classifying the held-out sample in the test data. We repeat the procedure on every sample and report the overall test accuracy in Table 4. We measured the classification accuracy using the Area Under the receiver operator Curve (AUC)^7^ denoting the number of true positives, true negatives, false positives and false negatives as TP, TN, FP, and FN respectively. AUC measures the area under the receiver operating characteristic curve which plots the true positive rate (TPR) and false positive rate (FPR) at different classification thresholds.

## Results

100 patients with matched GFAP and UCH-L1 concentrations were analyzed, 35 within the trauma with a head CT positive for vascular injury group, 10 within the spontaneous hemorrhage (SpontHem) group, 6 within the oxygen deprivation due to cardiac arrest or respiratory arrest (CA/RA) group, 10 within the computed tomography negative -high velocity trauma (CTN-HVT) group, and 39 within the control group (table 1). Log-transformation was performed on the concentrations of GFAP and UCH-L1 before conducting data analyses. Data collection was continuous except for the exclusion of one patient, who sustained a cardiac arrest resulting in a high speed collision and thus had a high velocity trauma confounded with an anoxic injury.

**Table 1:** Study demographics of patient groups stratified by injury type.

| Characteristic | Total | Traumatic Hemorrhage | CT (-) High Velocity Trauma | Spontaneous Hemorrhage | Cardiac/Respiratory Arrest | Healthy Control |
| --- | --- | --- | --- | --- | --- | --- |
| Subjects (n) | 100 | 35 | 10 | 10 | 6 | 39 |
| Age in years (range, mean) | 16-87, 46.5 | 22-85, 53.3 | 17-77, 42.7 | 45-80, 59.9 | 27-87, 51.5 | 16-65, 37.1 |
| Sex |  |  |  |  |  |  |
| Female | 45 (45%) | 9 (26%) | 2 (20%) | 3 (30%) | 2 (33%) | 29 (74%) |
| Male | 55 (55%) | 26 (74%) | 8 (80%) | 7 (70%) | 4 (67%) | 10 (26%) |
| Race |  |  |  |  |  |  |
| African American | 22 (22%) | - | 2 (20%) | 3 (30%) | 1 (17%) | 16 (41%) |
| Asian | 1 (1%) | - | 1 (10%) | - | - | - |
| Caucasian | 61 (61%) | 28 (80%) | 7 (70%) | 5 (50%) | 4 (67%) | 17 (44%) |
| Hispanic or Latino | 7 (7%) | 2 (6%) | - | 1 (10%) | - | 4 (10%) |
| Mixed Race | 1 (1%) | - | - | - | - | 1 (3%) |
| Native American | 3 (3%) | 2 (6%) | - | - | 1 (17%) | - |
| Unknown | 5 (5%) | 3 (9%) | - | 1 (10%) | - | 1 (3%) |
| Mechanism of Injury |  |  |  |  |  |  |
| Assault | 6 (6%) | 2 (6%) | 4 (40%) | - | - | - |
| Bicyclist Hit by Vehicle | 7 (7%) | 3 (11%) | 4 (40%) | - | - | - |
| Incidental Fall | 17 (17%) | 17 (47%) | - | - | - | - |
| Motor Vehicle Crash | 10 (10%) | 8 (22%) | 2 (20%) | - | - | - |
| Pedestrian Struck by Vehicle | 1 (1%) | 1 (3%) | - | - | - | - |
| Other | 20 (20%) | 4 (11%) | - | 10 (100%) | 6 (100%) | - |
| None | 39 (38%) | - | - | - | - | 39 (100%) |
| GCS on Arrival |  |  |  |  |  |  |
| 13 – 15 | 69 (69%) | 17 (48%) | 10 (100%) | 3 (30%) | - | 39 (100%) |
| 8 – 12 | 3 (3%) | 3 (9%) | - | - | - | - |
| 7 or less | 28 (28%) | 15 (43%) | - | 7 (70%) | 6 (100%) | - |
| Loss of Consciousness Duration |  |  |  |  |  |  |
| 0-30 mins | 10 (10%) | 8 (23%) | 2 (20%) | - | - | - |
| 30 mins – 24 hrs | 7 (7%) | 7 (20%) | - | - | - | - |
| Greater than 24 hrs | 5 (5%) | 1 (3%) | - | 3 (30%) | 1 (17%) | - |
| None | 52 (52%) | 5 (14%) | 5 (50%) | 3 (30%) | - | 39 (100%) |
| Unknown | 26 (26%) | 14 (40%) | 3 (30%) | 4 (40%) | 5 (84%) | - |

\*GCS: Glasgow Coma Scale

We first explored whether a single biomarker could separate the five different groups. The boxplot for each of the five groups is shown in Fig. 1. We observed a larger range in GFAP concentrations (Fig. 1A) compared to UCH-L1 (Fig. 1B). We performed one-way ANOVA analysis to test the null hypothesis that the mean is the same for all the five groups based on the GFAP and UCH-L1 concentrations, respectively. Next, we performed t-test and Wilcoxon rank-sum test on each of the ten combinations of the five groups to test whether using a single marker can distinguish each pair of groups. The p-values are shown in Table 2 and Table 3. Comparing with the controls, every one of the four subject groups has significantly different means (p < 0.01; t-test) and medians (p < 0·01; rank-sum test) on either GFAP or UCH-L1 concentrations.

**Table 2.** Comparison between types of injury: p-values of t-test (GFAP/UCH-L1)

|  | **SpontHem** | **CA/RA** | **CTN-HVT** | **Control** |
| --- | --- | --- | --- | --- |
| **Trauma** | 3·98E-03/2·96E-02 | 4·80E-06/4·35E-05 | 7·53E-12/5·44E-04 | 1·05E-30/2·21E-16 |
| **SpontHem** |  | 1·40E-06/1·25E-07 | 5·64E-12/1·29E-01 | 4·05E-26/5·49E-09 |
| **CA/RA** |  |  | 1·04E-01/1·15E-09 | 1·01E-07/2·55E-22 |
| **CTN-HVT** |  |  |  | 8·04E-08/7·80E-06 |

**Table 3.** Comparison between types of injury: p-values of Wilcoxon rank-sum test (GFAP/UCH-L1)

|  | SpontHem | CA/RA | CTN-HVT | Control |
| --- | --- | --- | --- | --- |
| Trauma | 2.88E-03/2.16E-02 | 2.57E-05/6.97E-05 | 2.81E-09/4.09E-04 | 4.92E-17/2.45E-13 |
| SpontHem |  | 1.14E-04/9.61E-06 | 4.79E-07/1.17E-01 | 1.77E-09/1.90E-06 |
| CA/RA |  |  | 3.51E-01/5.48E-06 | 6.58E-07/3.85E-08 |
| CTN-HVT |  |  |  | 3.76E-06/9.17E-05 |

**Figure 1.**
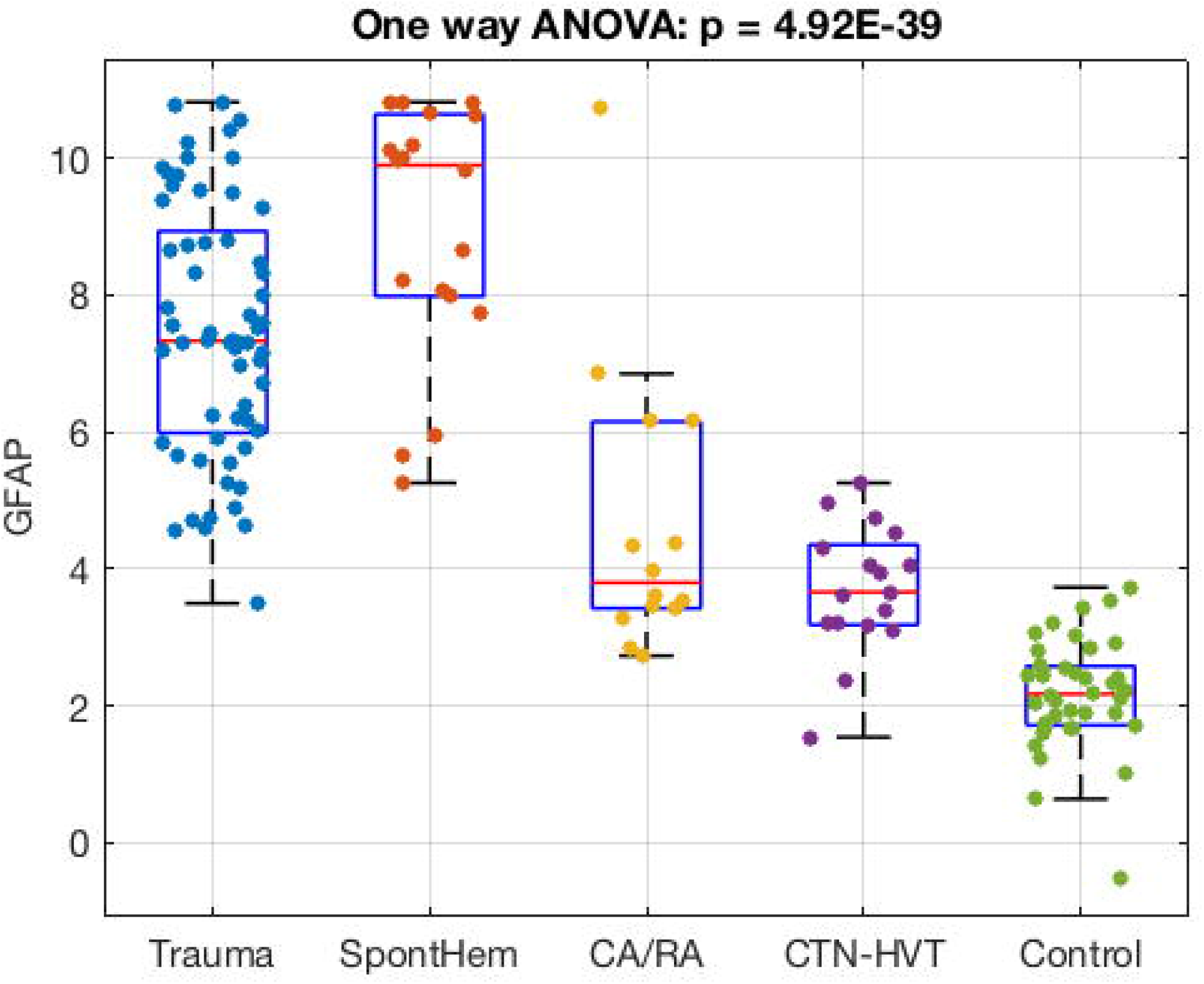
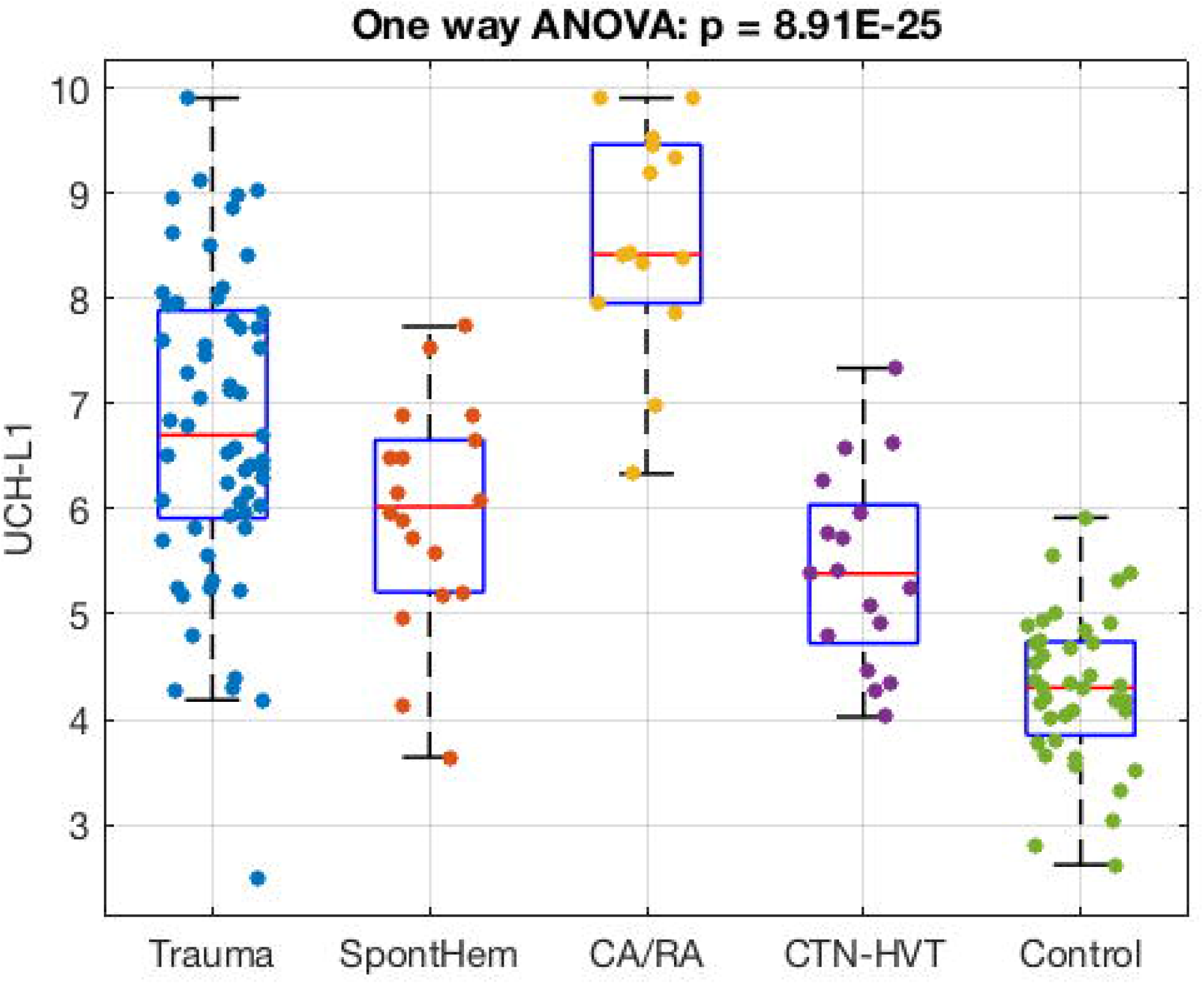
Box plots depicting serum marker concentrations of all blood draws taken from the five patient groups assessed by only a single biomarker. **A**. GFAP concentrations (log-transformed pg/mL) depict differences between brain injury groups: Trauma (blue), Spontaneous Hemorrhage (SpontHem shown in red), Cardiac or Respiratory Arrest (CA/RA shown in yellow), and CT-negative with high-velocity trauma (CTN-HVT shown in purple). **B**. UCH-L1 concentrations (log-transformed pg/mL) from patients with matching blood draw time points from GFAP concentrations. Significance shown in the plot title by P-value for one-way ANOVA analysis for each separate biomarker to stratify patient groups by injury. GFAP and UCH-L1 concentrations were transformed and log normalized.

We observed that samples in CTN-HVT can be distinguished from controls using either GFAP or UCH-L1. We performed t-test and Wilcoxon rank-sum tests to test how significant the difference on each single biomarker is between the two groups. The small p values shown in Tables 2 and 3 suggest the samples of CTN-HVT have significantly different means and medians.

**Table 4.** Classification with Support Vector Machine (SVM) based on the combinations of UCH-L1 & GFAP Area Under the Curve (AUC)

|  | SpontHem | CA/RA | CTN-HVT | Control |
| --- | --- | --- | --- | --- |
| Trauma | 0.83 (0.71, 0.95) | 0.95 (0.88, 1.00) | 0.97 (0.92, 1.00) | 1.00 |
| SpontHem |  | 1.00 | 1.00 | 1.00 |
| CA/RA |  |  | 0.98 (0.91, 1.00) | 1.00 |
| CTN-HVT |  |  |  | 0.93 (0.81, 1.00) |

We explored whether the GFAP and UCH-L1 concentrations can be combined as predictors to classify the samples in each pair of the five different groups using machine learning with the Support Vector Machine (SVM). The concentrations of GFAP and UCH-L1 for each patient sample within the four subject groups and controls were plotted in Fig. 2. We measured the classification accuracy with the Area Under the receiver operator Curve (AUC) scores which are shown in Table 4. One can observe that the subjects with trauma, SpontHem and CR/RA could be distinguished from controls with AUC = 1·00. In the comparisons within the four subject groups, the AUCs are all larger than 0·95. The high AUC of 0·93 suggest that the biomarkers GFAP and UCH-L1 can jointly distinguish the CTN-HVT samples from controls.

**Figure 2.**
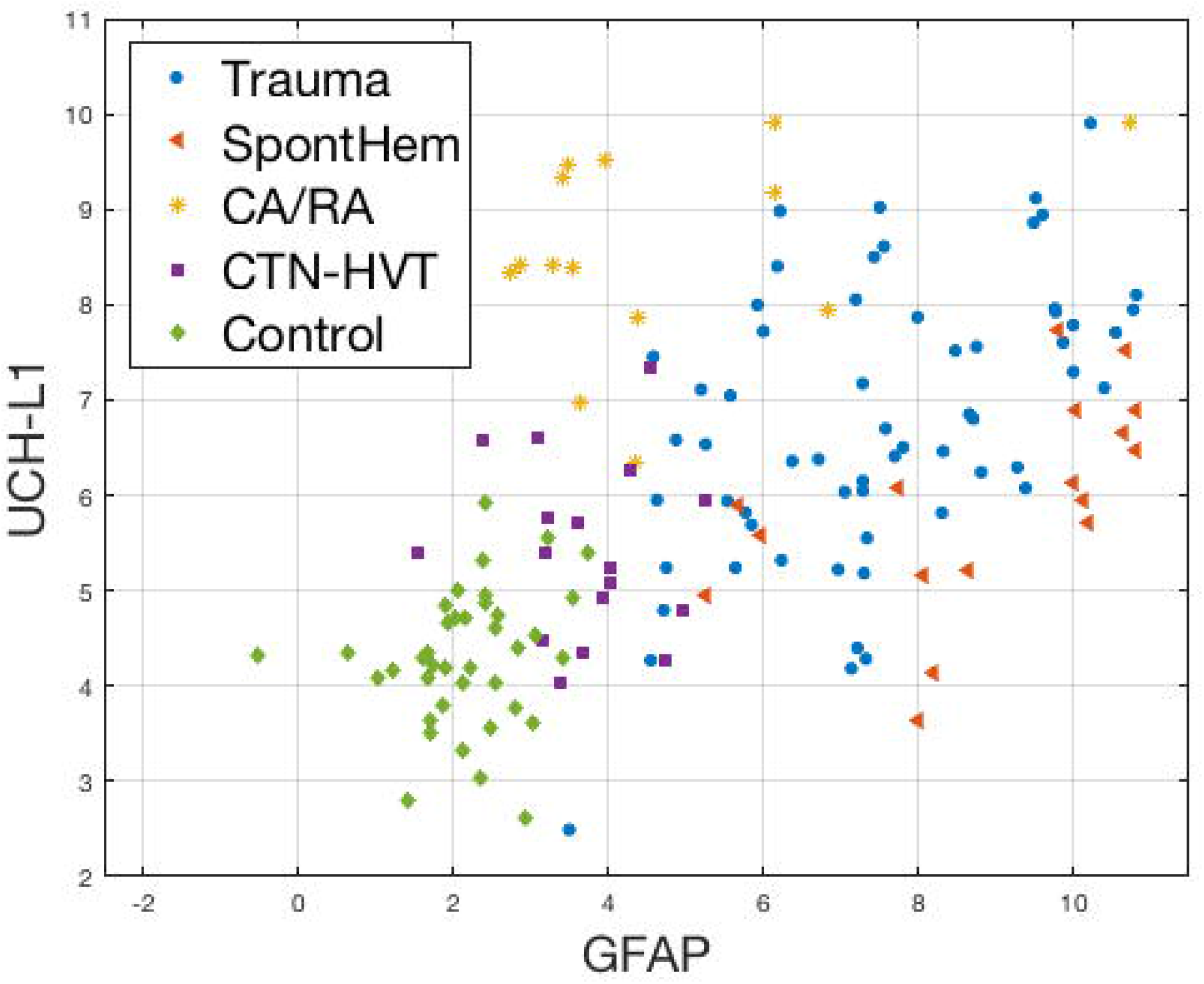
Scatter plot of GFAP and UCH-L1 concentrations from all patient samples show the four separate concentration profiles used by the SVM to predict etiology in comparison to the uninjured control samples. Uninjured controls (shown as green diamonds), Traumatic injury (shown as blue circles), Spontaneous hemorrhage (SpontHem show as red triangles), Oxygen deprivation from cardiac or respiratory arrest (CA/RA shown as yellow asterisks), and computed tomography negative with high-velocity trauma (CTN-HVT shown as purple squares). Log-transformed pg/mL concentrations are shown on scatter plot axes.

We next explored whether the lesion volumes derived from the CT scan images together with the GFAP and UCH-L1 concentrations can be combined as predictors to classify the samples in each pair of the five different groups using SVM. We measured the classification accuracy with the receiver operator curve (AUC) scores which are shown in Table 5. We observe that including the lesion volumes as additional predictors can significantly improve the classification accuracy.

**Table 5.** Classification with Support Vector Machine SVM based on the combinations of serum biomarkers and CT images Area Under the Curve (AUC)

|  | **SpontHem** | **CA/RA** | **CTN-HVT** | **Control** |
| --- | --- | --- | --- | --- |
| **Trauma** | 0.96 (0.91, 1.00) | 0.99 (0.96, 1.00) | 0.98 (0.94, 1.00) | 1.00 |
| **SpontHem** |  | 1.00 | 1.00 | 1.00 |
| **CA/RA** |  |  | 0.98 (0.91, 1.00) | 1.00 |
| **CTN-HVT** |  |  |  | 1.00 |

## Discussion

We evaluated serum biomarker concentrations of ubiquitin c-terminal hydrolase-L1 (UCH-L1) and glial fibrillary acidic protein (GFAP) in the acute period of traumatic brain injury (<32 hours of injury) to differentiate between five different groups of patients: uninjured controls, CT- negatie injury with high-velocity trauma (inertial injury), oxygen deprivation injury (CA/RA), spontaneous hemorrhage (non traumatic injury) and CT-positive vascular traumatic injury. Evaluation of two serum biomarkers together was able to differentiate between traumatic injuries and non traumatic injuries, due to a range of causes. Biomarker profiles differ amongst the groups because the mechanisms through which the biomarkers are being released and to what magnitude varies. Oxygen deprivation led to the highest concentrations of UCH-L1 across all types of injuries examined and had lower levels of GFAP compared to the trauma and spontaneous hemorrhage groups.

GFAP and UCH-L1 serum levels measured immediately after a traumatic injury correlated with increased likelihood of poor outcomes six months post injury. Previously, GFAP alone was able to accurately predict traumatic injuries and no improvement was found by adding UCH-L1 concentrations to the analysis.^8^ GFAP and UCHL-1 concentrations were also assessed in patients with mild TBI. These biomarkers were increased in patients with unfavorable short-term outcomes. These levels were significant within 6 hours post injury.^9^

Concentrations of GFAP were more elevated for spontaneous hemorrhage as compared to injuries caused by trauma. Conversely, UCH-L1 concentrations were modestly increased for trauma-induced injuries in comparison to spontaneously presenting hemorrhages. Increased GFAP concentrations may also be due to larger volumes of blood found in spontaneous hemorrhages. Patients who experience multiple TBIs may potentially develop autoantibodies against GFAP that may cause reduced levels of GFAP with subsequent injuries.^10^ Thus we would caution against use of a single marker in classifying injury.

Consistent with previous studies, we were able to accurately predict an ischemic stroke from hemorrhagic stroke, where immediate cell death results in a rise in GFAP concentration in individuals who experienced a hemorrhagic stroke.^11^ GFAP levels from ischemic patients were still significantly elevated compared to healthy controls. However, we found robust serum levels of UCH-L1 from ischemic injury, enabling better classification, when both biomarkers are combined. We demonstrate improved predictive potential using both UCH-L1 and GFAP as shown in Figure 2 with AUC of 0·93. The AUC values suggest a reasonable model to classify brain injury.

Addition of an open source, automated CT scan analysis (BLAST CT) to aid in stratification of injury by etiology resulted in additional improvement in differentiating traumatic injuries from those occurring from a spontaneous hemorrhage (AUC of 0.83 in table 1 and 0.96 in table 2). BLAST-CT identifies three types of hemorrhage, which when assessed in conjunction with concentration of the two serum biomarkers further improved the algorithm (table 5). These improvements most likely are due to the inclusion of lesion volume as well as identification of specific hemorrhage type.

In this study, we focused on blood draws taken within 32 hours of presentation to the emergency department. Both biomarkers are known to have different kinetic properties due to their location and release within the body. UCH-L1 peaks early and decreases rapidly in intracranial lesions, and GFAP increases after 4 hours and declines at 16 hours.^12^ Previously, GFAP and UCH-L1 concentrations acquired within 48 hours post injury were capable of distinguishing mass lesions from diffuse injuries. Analysis of concentrations up to seven days post injury did not increase predictive power to differentiate between injury type.^13^ A major challenge for diagnosing traumatic brain injuries is that patients can present with a wide range of symptoms and/or have altered consciousness due to other processes such as concomitant medications, intoxication, and comorbid injuries. Furthermore, some symptoms can be milder and harder to differentiate from other disorders, leading patients to delay presentation to the emergency department until they experience more severe symptoms. We found these two biomarkers, along with algorithmic CT analysis, to improve classification of injuries of different etiology with little dependence on the timing of the blood draw when taken within 32 hours of presentation to the emergency department.

## Conclusion

The findings of this study demonstrate the utility of using two serum biomarkers and algorithmic analysis of CT scan for classification of diverse brain injuries. This study is limited to small sample sizes for each type of pathology and necessitates investigation of larger sample sizes to overcome potential sampling bias. Our future goal is to perform clinical validation studies using the serum biomarker prediction algorithm developed in this smaller, developmental study on larger dataset such as the Transforming Research and Clinical Knowledge in Traumatic Brain Injury initiative (TRACK-TBI).^15^ Further, the predictive power of the biomarkers was only assessed by acute clinical outcomes from known etiologies. In future work, we will add additional objective markers, such as eye tracking, to enable assessment of severity in addition to type of injury. Information from these studies may also elucidate biological differences in race and sex that are known to exist across the spectrum of brain injuries. These diagnostic tools will improve the quality of life for patients, give guidance to families of the injured, and create objective measures for physicians.

## Data Availability

Data will be made available upon reasonable request.

https://tracktbi.ucsf.edu/transforming-research-and-clinical-knowledge-tbi

