## Supplemental Materials for "Machine Learning with Objective Serum Markers and Algorithmic Deep Learning Computed Tomography Scan Analysis for Classification of Brain Injury"

**Supplemental Methods**

Participants included trauma patients of all ages presenting to a single hospital Emergency Department (ED), trauma bay, or as direct transfer to neurosurgery between May 2016 and April 2017. Patients were excluded if they had a major psychiatric or neurological disorder, developmentally abnormal, or were prisoners. At the time of admission, potential participants underwent screening before providing informed consent. This screening process is necessary, because many of the patients admitted with trauma were in need of time-sensitive care that would become compromised by the time it takes to obtain informed consent for this minimal risk study. Informed consent was obtained upon completion of the screening process as soon as it became appropriate due to either (1) available time, (2) recovery of cognitive competence, or (3) the presence of a legal proxy. In the case that a potential participant died before informed consent was obtained, and no legal proxy was determined, consent to participate in the study was waived. These procedures have been assessed by the appropriate review committee. Patient information was identified by searching EPIC medical records for all trauma admissions and cross-checking with the American College of Surgeons trauma registry utilized in the hospital. 779 brain injury subjects and healthy controls enrolled during the study period and 331 were determined to have at least one valid biomarker result within 32 hours of injury. Clinical diagnosis and assignment to brain injury groups was performed by trained clinicians, based on clinical findings. CT scans were acquired as part of standard care. The Glasgow Coma Scale and the duration of loss of consciousness were two variables taken upon admission to hospital and reported in this study.

Blood collection was obtained at the time of admission for clinical purposes, and additional specimen were obtained and retained for research purposes. This study focused on a subset of blood draws taken as part of the CLASSIFY clinical trial (NCT02706574). Patients had up to three blood draws taken within 32 hours of hospital admission. Study participants contributed at least one and up to three blood draws with target times of 0 (as close to ictus/injury occurrence as feasible), 3, and 24 hours. GFAP and UCHL1 concentrations from all available time points were matched and utilized for data analysis. These three time points were assessed to determine the kinetic changes over time as we previously demonstrated [4]. For the development of the machine learning algorithm capable of stratification by injury etiology, all available serum biomarker concentrations taken from the same blood draw the were utilized. Due to the nature of traumatic injuries many patients only had one or two blood draws taken. There was no correlation between injury etiology and number of blood draws taken (Supplemental Table 1). Uninjured (healthy) control patients had one blood draw taken. The blood was drawn through venipuncture or via a central venous access if one was required by the standard of care. Blood draws were taken within 32 hours of presentation to the hospital. UCH-L1 and GFAP serum concentrations were measured using the recently FDA-authorized i-STAT system (Abbott).

### Machine Learning

Support vector machine and kernels:

Given a set of training data $\{(\mathbf{x}_i, y_i)\}_{i=1}^N$ with $\mathbf{x}_i$ be the $i$-th sample vector and $y_i \in \{+1, -1\}$ as its label, the SVM outputs the score $f(\mathbf{x}_{\text{new}})$ of a new sample $\mathbf{x}_{\text{new}}$ as

$$f(\mathbf{x}_{\text{new}}) = \sum_{i=1}^{N} \alpha_i y_i K(\mathbf{x}_i, \mathbf{x}_{\text{new}}) + b, \tag{1}$$

where $K(\mathbf{x}_i, \mathbf{x}_j)$ is the kernel function defined with a mapping of the original feature space into a transformed space for non-linear classification. We use the Gaussian kernel defined below as the non-linear kernel function:

$$K(\mathbf{x}_i, \mathbf{x}_j) = \exp(-\frac{||\mathbf{x}_i - \mathbf{x}_j||^2}{\sigma^2}) \tag{2}$$

The $\alpha_i$ for $i = 1, \ldots, N$ are obtained by solving the optimization problem

$$\underset{\alpha_1,\ldots,\alpha_N}{\text{maximize}} \sum_i \alpha_i - \frac{1}{2} \sum_{i,j=1}^{N} \alpha_i \alpha_j y_i y_j K(\mathbf{x}_i, \mathbf{x}_j) \tag{3}$$

$$\text{subject to } \sum_{i=1}^{N} \alpha_i y_i = 0, \text{ and } 0 \leq \alpha_i \leq \frac{1}{2N\lambda}, \ \forall i = 1, \ldots, N, \tag{4}$$

where $\lambda$ is a hyper-parameter.

After obtaining all the $\alpha_i$s, the scalar $b$ is obtained as:

$$b = -0.5 * (\max_{j:y_j=-1} \sum_{i=1}^{N} \alpha_i y_i K(\mathbf{x}_i, \mathbf{x}_j) + \min_{j:y_j=1} \sum_{i=1}^{N} \alpha_i y_i K(\mathbf{x}_i, \mathbf{x}_j)). \tag{5}$$
