## Supplemental Table 1 for "Machine Learning with Objective Serum Markers and Algorithmic Deep Learning Computed Tomography Scan Analysis for Classification of Brain Injury"

**Supplemental Table 1** -Number of blood draws per etiology

| | Trauma | SpontHem | CA/RA | CTN-HVT | Control |
|---|---|---|---|---|---|
| 1 blood draw | 19 | 5 | 0 | 4 | 39 |
| 2 blood draw | 6 | 2 | 4 | 5 | 0 |
| 3 blood draw | 10 | 3 | 2 | 1 | 0 |
